# From genes to pathways: genetic convergence in early-onset Parkinson’s disease in India

**DOI:** 10.64898/2026.08.31.26361762

**Authors:** Ramesh Menon, Aayat Ibrahim Khan, Dayanandhi Elangovan, Rukmini Mridula Kandadai, Vinay Goyal, Soaham Dilip Desai, Deepika Joshi, Hrishikesh Kumar, Pettarusup M Wadia, Adreesh Mukherjee, Niraj Kumar, Sahil Mehta, Thenral S Geetha, Sandeep Chargulla, Sakthivel Murugan, Malini Venkata, Heli S Shah, Vijayshankar Paramanandam, Mitesh Chandarana, Ravi Yadav, Rajinder K Dhamija, Pramod Kumar Pal, Atanu Biswas, Ravi Gupta, Rupam Borgohain, Vedam Ramprasad, Prashanth Lingappa Kukkle, Parkinson Research Alliance of India (PRAI)

## Abstract

Parkinson’s disease (PD) arises through disruption of multiple interconnected cellular processes, but the genetic contributions to these processes may differ across ancestries. We investigated functional convergence among genes harboring pathogenic or likely pathogenic (P/LP) variants and variants of uncertain significance (VUS) in a multicenter Indian cohort recruited through the Genetics of Parkinson’s Disease in India–Young-Onset Parkinson’s Disease project (GOPI-YOPD).

The cohort included 668 participants (463 males-69.3%) with a mean age at motor onset of 39.4±8.8 years. P/LP variants and VUS identified through previously reported whole-exome or whole-genome sequencing were retained as separate evidential categories. The P/LP-associated gene set comprised 11 unique genes and the VUS-associated set comprised 40 unique genes. Separate STRING functional-enrichment analyses evaluated Gene Ontology Biological Process, Molecular Function and Cellular Component terms, KEGG pathways, WikiPathways and STRING local-network clusters. Terms meeting a Benjamini–Hochberg false-discovery-rate threshold of <0.05 were organized into eight non-mutually-exclusive ontology/pathway categories. Gene-to-pathway mappings were subsequently projected to individual participants to estimate pathway representation and examine clinical associations.

At least one reportable P/LP variant or VUS was identified in 336/668 participants (50.3%): 35 had a P/LP variant alone, 282 had VUS alone and 19 had a P/LP variant together with VUS in one or more additional genes. The most frequently represented categories were mitochondrial organization (247/336, 73.5%), autophagy-related processes (228/336, 67.9%) and regulation of synaptic-vesicle transport (201/336, 59.8%). *PRKN* was the most frequent P/LP-associated gene, occurring in 29/54 P/LP carriers, followed by *PLA2G6* and *PINK1*. Lysosomal transport was represented exclusively by VUS-associated genes, particularly *GBA1*, *VPS13C* and *LRRK2*. Among P/LP carriers, additional VUS in distinct genes were not associated with age at onset (P = 0.81) or family history (52.6% versus 31.4%; P = 0.15). No pathway–phenotype association remained significant after correction for multiple testing.

Genetic findings in this Indian cohort converged across an interconnected mitochondrial–autophagic–lysosomal–vesicular network, with different contributions from P/LP-associated and VUS-associated gene sets. This study provides the first pathway-resolved South Asian genetic profile and a framework for comparative studies across populations.

## Introduction

Parkinson’s disease (PD) arises through disruption of multiple interconnected cellular processes, including mitochondrial quality control, autophagy–lysosomal degradation, vesicular and endosomal trafficking, synaptic function, lipid metabolism, proteostasis and neuroimmune regulation.^1^ Its genetic architecture similarly extends from rare, highly penetrant variants to low-frequency and common risk alleles. Variants distributed across different genes may therefore converge on the same biological pathway, while cumulative perturbations involving more than one pathway may contribute to disease susceptibility and clinical heterogeneity.^2^

The genetic architecture of PD is not uniform across populations.^3^ Differences in the frequency, spectrum and effect size of variants in genes such as GBA1, LRRK2, PRKN and PINK1 are increasingly recognized across European, African, Latin American and Asian ancestries.^4^ However, most studies have examined individual genes, selected variants or a single predefined pathway, frequently in populations of predominantly European ancestry.^5^ Consequently, it remains uncertain whether the relative genetic load across major PD-related pathways differs between populations. This distinction is relevant because a shared clinical phenotype may arise through different molecular routes, with implications for genetic interpretation, biological stratification and the development of pathway-directed therapies.

Young-onset PD, in which genetic contributions are more readily identifiable, provides an informative setting to examine this question. India remains underrepresented in global PD genomic studies despite its considerable genetic diversity and substantial young-onset PD population.^3,6–8^ In the present study, we examined patient-level P/LP and VUS findings from a multicentre Indian cohort enriched for early-onset PD, evaluated their functional convergence across biological pathways, and mapped these pathways back to individual participants to explore their clinical and demographic correlates. We aimed to define the pathway-resolved genetic profile of Indian early-onset PD and to determine whether co-occurring variants were associated with phenotypic variation.

## Materials and Methods

### Study cohort and variant classification

This secondary analysis used patient-level clinical and genetic data from the multicentre Genetics of Parkinson’s Disease in India–Young-Onset Parkinson’s Disease (GOPI-YOPD) project. The clinical characteristics and genetic findings of this cohort have been reported previously.^93,6^ Recruitment primarily targeted patients with Parkinson’s disease (PD) and an age at onset (AAO) of motor symptoms ≤50 years from specialist movement-disorder centers across India. The source dataset additionally included affected later-onset relatives recruited through qualifying families.^3^

Recruitment, clinical assessment, whole-exome or whole-genome sequencing, variant calling and American College of Medical Genetics and Genomics/Association for Molecular Pathology–based variant classification were performed as previously described.^10^

Variants previously classified as pathogenic or likely pathogenic (P/LP) and variants of uncertain significance (VUS) were retrieved with their corresponding genes and patient-level clinical data. These were retained as separate evidential categories throughout the analysis. A VUS was not considered evidence of disease causality, and its assignment to a pathway represented gene-level functional annotation rather than demonstrated pathway dysfunction.

### Gene-set enrichment and pathway assignment

The P/LP gene set comprised 11 unique genes harbouring at least one P/LP variant, and the VUS gene set comprised 40 unique genes harbouring at least one VUS. A gene was included in both sets when P/LP and VUS findings occurred in different participants. Each gene was counted once within each set, irrespective of the number of variants or affected participants. The analysis therefore evaluated functional convergence among implicated genes rather than patient-frequency-weighted or case– control variant burden.

Functional enrichment was performed separately for the two gene sets using STRING v.12.0, 06/07/2026, with *Homo sapiens* as the reference organism.^11^ Gene Ontology Biological Process, Molecular Function and Cellular Component terms, KEGG pathways and WikiPathways were evaluated. Enrichment was tested using STRING’s hypergeometric test with Benjamini–Hochberg false-discovery-rate (FDR) correction; terms with FDR<0.05 were considered significant. Enrichment strength was expressed as log_₁₀_(observed/expected gene count).

Enriched terms were matched between the P/LP and VUS outputs using exact ontology or pathway identifiers and classified as shared or variant-class-only. Biologically overlapping terms were consolidated into eight non-mutually-exclusive pathway domains. A pathway was considered represented in an individual when at least one retained P/LP variant or VUS occurred in a gene assigned to that pathway. Because genes and participants could map to multiple pathways, pathway percentages were not expected to sum to 100%. Complete STRING outputs, term-selection procedures and gene-to-pathway mappings are provided in the Supplementary Table 1.

### Clinical and statistical analysis

Participants were classified as P/LP-only, VUS-only, P/LP plus an additional VUS in a distinct gene, or without a reportable P/LP/VUS finding. The primary comparison was between P/LP-only and P/LP-plus-additional-VUS carriers; VUS-only single-gene and multigene carriers were compared separately as a negative comparator. Continuous variables were summarized as median (interquartile range) and compared using the Mann–Whitney U or Kruskal–Wallis test, with Holm-adjusted Dunn tests following significant multi-group comparisons. Categorical variables were compared using chi-square or Fisher’s exact tests. Pathway–phenotype comparisons were exploratory and were corrected collectively using the Benjamini–Hochberg’s FDR method. Analyses used available cases without imputation, with non-missing denominators reported. All tests were two-sided, effect estimates were accompanied by 95% confidence intervals were estimable, and adjusted p<0.05 was considered significant. Analyses were conducted using Excel MSO 2021 and R (Ver.4.5.3).

### Ethics approval

The GOPI-YOPD project was approved by the institutional ethics committee at each participating centre. Written informed consent was obtained from all participants.

### Data Availability statement

De-identified data underlying this study may be made available by the GOPI-YOPD Consortium upon reasonable request to the corresponding author, subject to approval by the consortium’s data-access committee and applicable ethical, consent and data-sharing requirements.

## Results

### 1. Overview of the Cohort

The cohort comprised 668 participants with juvenile-, young- or early-onset PD recruited across India; 463 (69.3%) were male. The mean age at onset was 39.4±8.8 years, and the mean duration of motor symptoms was 92.3±65.8 months. Among participants assignable to the prespecified onset categories, 21 had juvenile-onset PD (JOPD; AAO≤20 years), 321 had young-onset PD (YOPD; AAO 21–40 years), and 300 had early-onset PD (EOPD; AAO 41–50 years). (Table-1) The cohort additionally included 26 affected relatives with AAO >50 years who were recruited through families containing a qualifying early-onset participant. (Table-1)

**Table 1:** Demographic, clinical and genetic characteristics of the study cohort, stratified by reportable genetic finding status and age-at-onset category.

At least one qualifying P/LP variant or VUS was identified in 336 participants (50.3%). These included 35 participants (5.2% of the complete cohort) with P/LP variants alone, 282 (42.2%) with VUS alone, and 19 (2.8%) with a P/LP variant accompanied by VUS in one or more additional genes. The latter multiple-hit group comprised 13 dual-hit and six triple-hit participants. Overall, 54 participants (8.1%) carried at least one P/LP variant, and 301 (45.1%) carried at least one VUS. (Figure-1)

**Figure 1.**
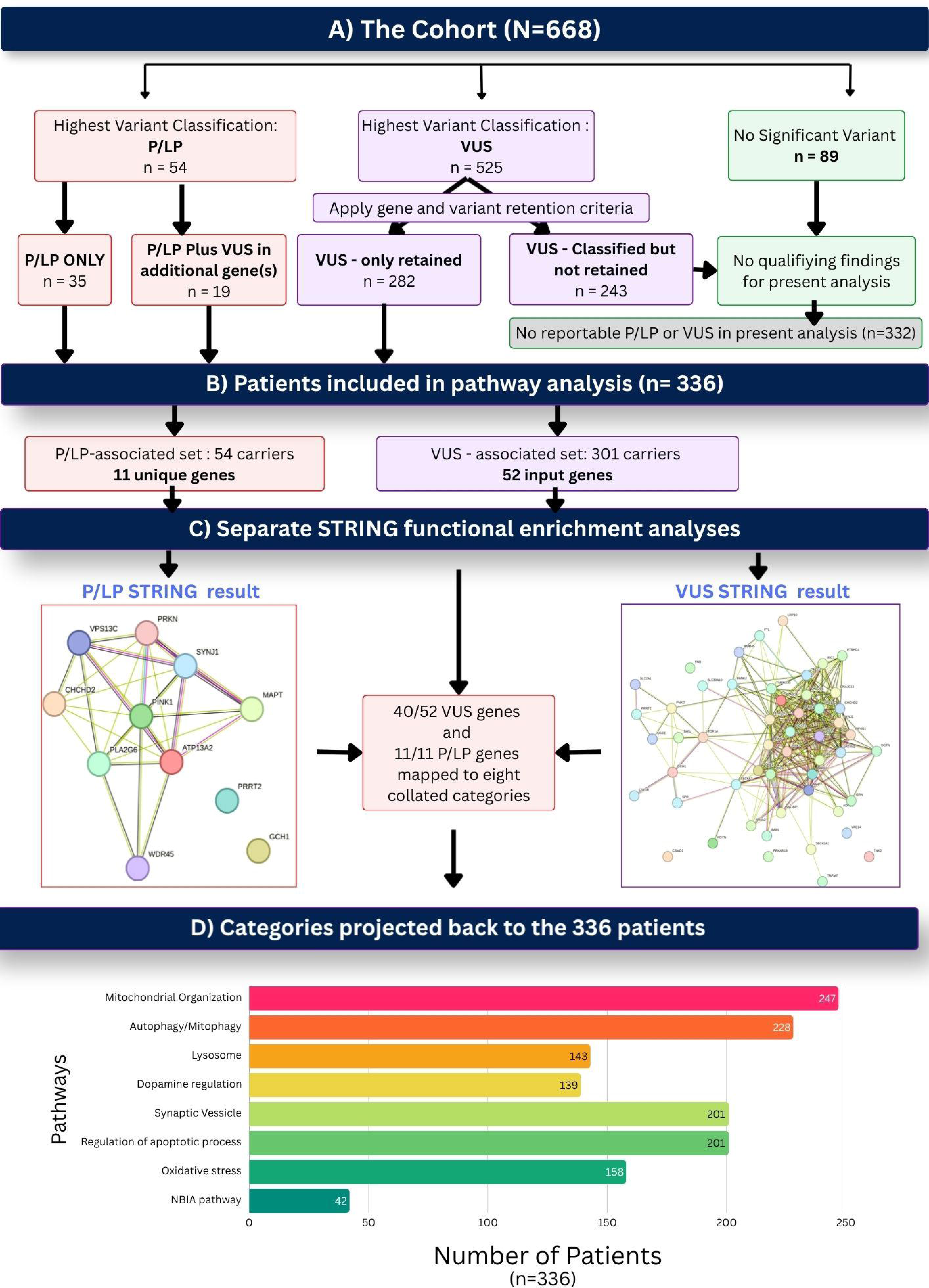
Study workflow from source variant classification to patient-level pathway representation `o` **(A)** The source cohort comprised 668 participants, assigned to mutually exclusive groups according to their highest variant classification: pathogenic/likely pathogenic (P/LP; n = 54), variant of uncertain significance (VUS; n = 525), or no significant variant (n = 89). The P/LP group included participants who also carried VUS. Following application of the gene- and variant-retention criteria for the present analysis, 35 participants had P/LP variants alone, 19 had a P/LP variant together with VUS in one or more additional genes, and 282 had retained VUS without a P/LP finding. These 336 participants were included in the patient-level pathway analysis. Of the 525 participants whose highest source classification was VUS, 243 did not have a VUS meeting the present retention criteria; together with the 89 participants with no significant variant, they constituted the 332 participants without a qualifying retained finding. `o` **(B)** The 54 P/LP carriers contributed 11 unique genes to the P/LP gene set. The VUS gene set comprised 52 unique input genes identified among 301 carriers, including 282 VUS-only participants and 19 participants with a P/LP variant plus additional VUS. Each gene was counted once within each variant-class gene set, irrespective of the number of variants or participants carrying findings in that gene. `o` **(C)** The P/LP and VUS gene sets underwent separate STRING functional-enrichment analyses incorporating Gene Ontology Biological Process, Molecular Function and Cellular Component terms, KEGG pathways, WikiPathways and STRING local-network clusters. Significance was assessed using Benjamini–Hochberg false-discovery-rate correction, with FDR < 0.05 considered significant. In the network visualizations, nodes represent proteins encoded by the submitted genes and edges represent STRING-supported functional associations. All 11 P/LP genes and 40 of the 52 VUS input genes contributed to the eight collated ontology/pathway categories. `o` **(D)** The eight non-mutually-exclusive categories were projected back to the 336 participants with qualifying findings. Patient-level representation denotes the number of participants carrying at least one retained P/LP variant or VUS in a gene assigned to the corresponding category. Because individual genes and participants could map to more than one category, category counts and percentages do not sum to the total cohort size or to 100%. Assignment of a VUS-containing gene to a category represents gene-level functional annotation and does not establish variant pathogenicity or demonstrate pathway dysfunction in the individual participant. `o` **Abbreviations:** FDR, false discovery rate; GO, Gene Ontology; KEGG, Kyoto Encyclopedia of Genes and Genomes; NBIA, neurodegeneration with brain iron accumulation; P/LP, pathogenic or likely pathogenic; STRING, Search Tool for the Retrieval of Interacting Genes/Proteins; VUS, variant of uncertain significance.

**Figure 2.**
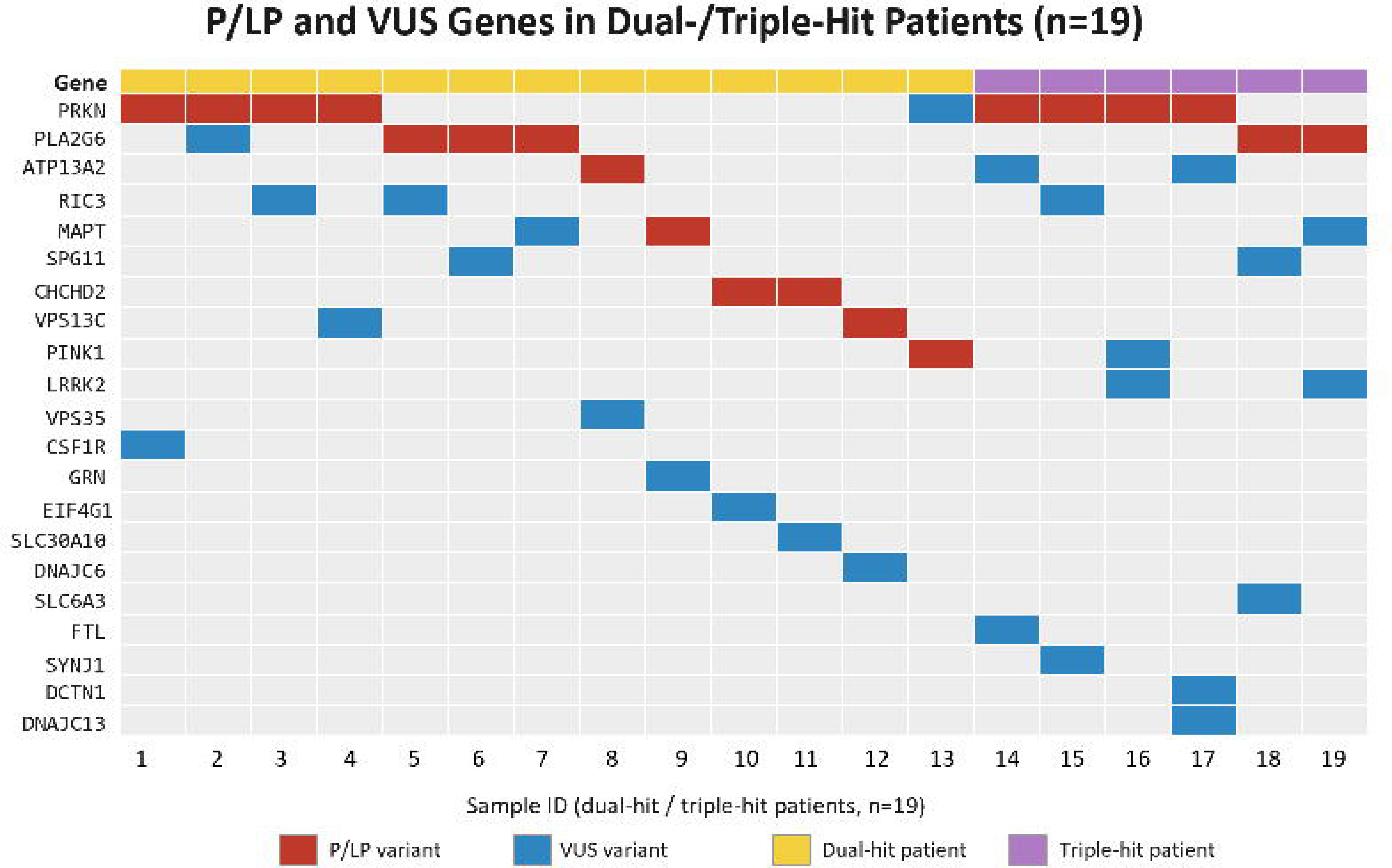
Gene-level co-occurrence of pathogenic/likely pathogenic variants and variants of uncertain significance among 19 participants. `o` Columns represent anonymized participants and rows represent genes harbouring at least one retained variant. Red cells indicate the primary gene containing a pathogenic or likely pathogenic (P/LP) variant, blue cells indicate an additional gene containing a variant of uncertain significance (VUS), and grey cells indicate that no retained variant was identified in that gene. The upper annotation bar distinguishes participants with a P/LP variant and VUS in one additional gene (yellow; n=13) from those with a P/LP variant and VUS in two or more additional genes (purple; n=6). The figure illustrates gene-level co-occurrence and does not establish oligogenic inheritance or a modifying effect of the additional VUS.

**Figure 3:**
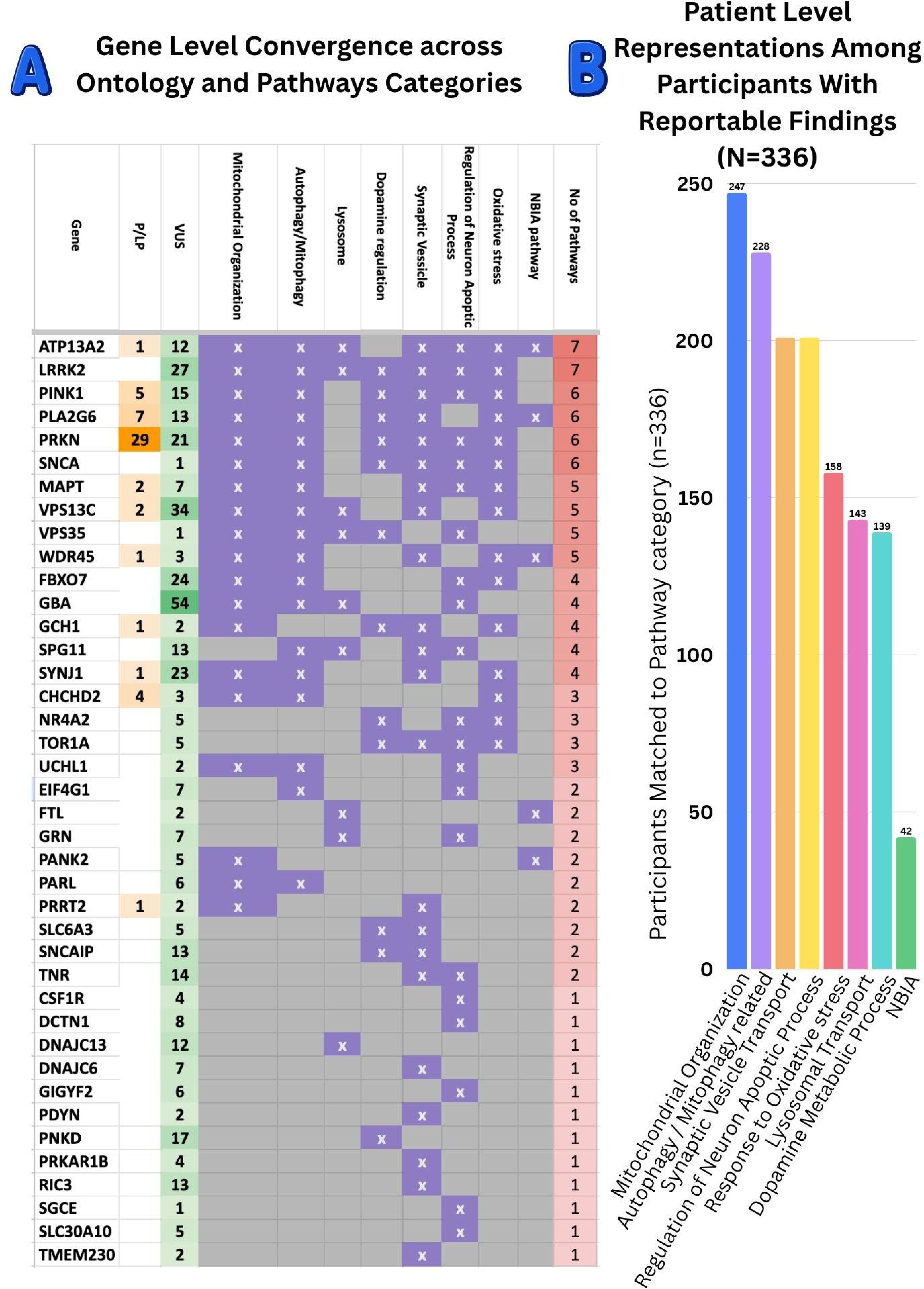
Genetic convergence across interconnected pathways in an Indian cohort enriched for early-onset Parkinson’s disease. `o` **(A)** Gene-level mapping of reportable findings across eight non-mutually-exclusive ontology and pathway categories. Rows represent genes, ordered by the number of pathway categories to which they were assigned. The first two columns show the numbers of participants with P/LP variants and VUS, respectively; filled matrix cells indicate assignment of a gene to the corresponding pathway category. **(B)** Patient-level representation of the eight categories among 336 participants with at least one reportable P/LP variant or VUS. Participants and genes could map to multiple categories; consequently, percentages do not sum to 100%. VUS mapping represents gene-level functional annotation and does not establish variant causality.

Compared with participants without a qualifying variant, those in the variant-detected group had a modestly earlier median AAO (39.0 versus 41.0 years; p=0.0487) and more frequently reported a family history of parkinsonism (24.1% versus 17.5%; p=0.0436). Sex distribution, consanguinity and dyskinesia frequency did not differ significantly between the groups (all p>0.36) (Table-1, Table-2).

**Table 2:** Patient-level representation and clinical characteristics across STRING-enriched ontology and pathway categories.

### 2. Functional enrichment and pathway representation

Functional enrichment analysis demonstrated convergence of the implicated genes across eight interrelated biological domains: mitochondrial organization, autophagy/mitophagy, lysosomal transport, dopamine metabolism, synaptic-vesicle trafficking, regulation of neuron apoptotic process, oxidative-stress response, and the neurodegeneration with brain iron accumulation (NBIA) pathway. The corresponding STRING, Gene Ontology and WikiPathways terms, contributing genes, enrichment strengths and FDR values are provided in Supplementary Table 1.

Patient-level pathway representation was determined by mapping each participant’s qualifying variants to these domains. Mitochondrial organization was the most frequently represented domain, involving 247/336 participants (73.5%), followed by autophagy/mitophagy in 228 (67.9%), synaptic-vesicle trafficking and regulation of neuron apoptotic process in 201 each (59.8%), oxidative-stress response in 158 (47.0%), lysosomal transport in 143 (42.6%), dopamine metabolism in 139 (41.4%), and the NBIA pathway in 42 (12.5%). Because individual genes could contribute to multiple biological processes and participants could carry variants in more than one gene, these groups overlapped and were not mutually exclusive.

Among the 54 participants carrying a P/LP variant, *PRKN* was the most frequently implicated gene (29/54, 53.7%), followed by *PLA2G6* (7/54, 13.0%) and *PINK1* (5/54, 9.3%). Among participants with VUS, *GBA1* was the most frequently represented gene (n=51), followed by *VPS13C* (n=33), *LRRK2* (n=26), and *FBXO7* (n=24). No P/LP variant in *LRRK2* was identified.

Two additional established areas of PD biology, α-synuclein-associated proteostasis and neuroinflammatory signalling did not meet the predefined threshold for retention as independent enriched domains. Proteostasis-related terms did not reach FDR <0.05 in the P/LP gene-set analysis, and only one participant carried an *SNCA* VUS. Neuroinflammatory signalling similarly did not form an independently enriched category; *CSF1R* VUS were identified in four participants. Failure to identify independent enrichment does not exclude the involvement of these mechanisms, which may overlap with the larger domains described above.

### 3. Pathway-specific findings

#### 3.1 Mitochondrial organization

Mitochondrial organization was represented in 247/336 participants (73.5%). P/LP representation was driven principally by *PRKN*, *PLA2G6*, *PINK1*, and *CHCHD2*, whereas the leading VUS-associated genes were *GBA1*, *VPS13C*, *LRRK2*, and *FBXO7*. The mean AAO among participants mapped to this domain was 37.8±9.8 years. Rest tremor was recorded in 186/244 participants with available data (76.2%) and was nominally more frequent than among participants not mapped to this domain (p=0.0189). No clear differences were observed in sex, polygenic risk score (PRS) category, consanguinity or dyskinesia frequency.

#### 3.2 Autophagy/mitophagy

The autophagy/mitophagy domain was represented in 228/336 participants (67.9%) and shared several contributing genes with mitochondrial organization, particularly *PRKN* and *PINK1*. Other prominent contributors included *PLA2G6*, *CHCHD2*, *GBA1*, *VPS13C*, *LRRK2*, *FBXO7*, *SPG11*, and *ATP13A2*. The mean AAO was 37.2±9.7 years. Notably, all 18 JOPD participants in the variant-detected cohort mapped to this domain, compared with none of the 108 participants outside it (nominal p=0.0061).

#### 3.3 Lysosomal transport

Lysosomal transport was represented in 143/336 participants (42.6%) and reached the predefined enrichment threshold in the VUS gene-set analysis but not in the P/LP gene-set analysis. The principal contributing genes were *GBA1*, *VPS13C*, *LRRK2*, *SPG11*, *ATP13A2*, and *DNAJC13*. The mean AAO was 39.1±8.4 years. Representation of this domain was more frequent among participants of East Indian origin than among participants not mapped to the domain (30.8% versus 16.1%; nominal p=0.0022). Nine participants carried a primary P/LP variant accompanied by VUS in one or more lysosomal or endolysosomal genes. The primary P/LP gene was *PRKN* in four participants and *PLA2G6* in three. Individual combinations are presented in Table 3.

**Table 3:** Comparison of clinical, demographic and genetic characteristics between participants carrying a pathogenic/likely pathogenic variant alone and those carrying a pathogenic/likely pathogenic variant plus one or more variants of uncertain significance in additional genes.

#### 3.4 Dopamine metabolism

The dopamine-metabolism domain was represented in 139/336 participants (41.4%). The principal P/LP-associated genes were *PRKN*, *PLA2G6*, *PINK1*, and *GCH1*, while the leading VUS-associated genes were *LRRK2*, *PRKN*, *PNKD*, *PINK1*, *SNCAIP*, and *SLC6A3*. The mean AAO was 37.3±10.6 years. JOPD was nominally more frequent among participants mapped to this domain than among non-carriers (9.4% versus 2.5%; p=0.0129), whereas constipation was less frequently reported (32.4% versus 47.4%; nominal p=0.0081).

#### 3.5 Synaptic-vesicle trafficking

Synaptic-vesicle trafficking was represented in 201/336 participants (59.8%). The leading contributors included *PRKN*, *PLA2G6*, *PINK1*, *LRRK2*, *SYNJ1*, *PNKD*, *SPG11*, *SNCAIP*, *TNR*, and *RIC3*. The mean AAO was 38.3±10.2 years. Dystonia was nominally more frequent among participants mapped to this domain than among those outside it (51/194, 26.3% versus 14.7%; p=0.0197).

All 12 participants with AAO >50 years in the variant-detected group mapped to the synaptic-vesicle-trafficking domain (6.0% of pathway carriers versus none of the participants outside the domain; nominal p=0.0096). These individuals represented the variant-detected subset of the familial LOPD participants retained through the recruitment strategy described above.

#### 3.6 Regulation of neuron apoptotic process

The regulation of neuron apoptotic process domain was represented in 201/336 participants (59.8%) and reached the enrichment threshold in the VUS gene-set analysis but not in the P/LP gene-set analysis. The principal contributors were *GBA1*, *LRRK2*, *FBXO7*, *PRKN*, *PINK1*, *SPG11*, *TNR*, and *ATP13A2*. The mean AAO was 39.2±9.1 years.

Reported consanguinity was lower among participants mapped to this domain than among those outside it (16/197, 8.1% versus 20.7%; nominal p=0.0014).

Representation was also nominally more frequent among participants of East Indian origin (26.4% versus 16.3%; p=0.0413). Differences across the individual YOPD and EOPD categories were of borderline nominal significance and are provided in Table 2.

### 3.7 Oxidative-stress response

The oxidative-stress-response domain was represented in 158/336 participants (47.0%). The principal contributing genes included *PRKN*, *PLA2G6*, *PINK1*, *CHCHD2*, *LRRK2*, *FBXO7*, and *ATP13A2*. The mean AAO was 36.8±10.5 years. This domain showed the strongest nominal association with JOPD: 16/158 participants mapped to the domain (10.1%) had AAO ≤20 years, compared with 2/178 participants outside it (1.1%; nominal p=0.0006).

### 3.8 NBIA pathway

The NBIA pathway was the least frequently represented domain, involving 42/336 participants (12.5%). P/LP-associated genes comprised *PLA2G6*, *ATP13A2*, and *WDR45*, while VUS-associated genes included *PLA2G6*, *ATP13A2*, *PANK2*, *WDR45*, and *FTL*. This group had the lowest mean AAO among the eight domains (34.8±12.4 years). JOPD was more concentrated among participants mapped to the NBIA domain than among those outside it (7/42, 16.7% versus 11/294, 3.7%; nominal p=0.0018). A nominal difference in sex distribution between multiple-hit and other NBIA-pathway participants was observed in small subgroups and is reported in Table 2.

### 4. Co-occurring variants across multiple genes

Among the 54 participants carrying a P/LP variant, 35 had no VUS in an additional gene, whereas 19 carried VUS in one or more additional, distinct genes. The latter represented 5.7% of participants with a qualifying genetic finding and 2.8% of the complete cohort. Thirteen participants carried a P/LP variant plus a VUS in one additional gene, and six carried a P/LP variant together with VUS in two or more additional genes.

In the direct comparison, AAO was similar between P/LP-only and P/LP-plus-additional-VUS carriers (mean 30.8±10.0 versus 30.5±10.4 years; p=0.81). Disease duration, sex distribution and consanguinity also did not differ significantly between the groups. Family history was numerically more frequent among P/LP-plus-additional-VUS carriers than among P/LP-only carriers (52.6% versus 31.4%), although this difference was not statistically significant (p=0.15). The distribution across juvenile-, young-and early-onset categories was also comparable. These findings did not support an association between the presence of an additional VUS and earlier AAO among participants already carrying a P/LP variant.

Among the 282 participants with VUS but no P/LP finding, 78 (27.7%) carried VUS in two or more genes and 204 carried a VUS in only one gene. Of the 78 VUS-only multigene carriers, 50 (64.1%) had variants in genes sharing at least one pathway domain, whereas 28 (35.9%) had no overlap among their assigned pathways. VUS-only multigene carriers did not have an earlier median AAO than single-gene VUS carriers (40.0 versus 41.0 years; p=0.600), and their family-history frequencies were comparable (23.1% versus 20.6%; p=0.927).

*PRKN* was the primary P/LP gene in 8/19 P/LP-plus-additional-VUS carriers, followed by *PLA2G6* in 5/19. Several co-occurring findings involved genes mapping to shared or biologically connected processes, including autophagy/mitophagy, mitochondrial quality control and lysosomal or endo-lysosomal trafficking. One participant carried a P/LP *PINK1* variant together with a *PRKN* VUS, involving two genes within the established PINK1–PRKN mitophagy axis. These combinations indicate potential biological convergence but, in the absence of segregation or functional evidence, were not interpreted as established digenic or oligogenic causation. Individual variants, gene combinations and pathway assignments are detailed in Table 3

### 5. Polygenic risk score and multiple-testing assessment

PRS classification was available for 314/336 participants (93.5%); 87 (27.7%) were classified as high PRS and 227 (72.3%) as low PRS. High-PRS status was not significantly associated with any of the eight pathway domains, P/LP-plus-additional-VUS status, or the evaluated motor and non-motor features (all nominal p≥0.10). No detectable relationship between the measured PRS category and pathway representation or multigene findings was therefore identified.

A total of 248 pathway–clinical-variable comparisons were performed. Eighteen (7.3%) yielded nominal p-values <0.05, but none remained significant after Bonferroni or Benjamini–Hochberg false-discovery-rate correction. Accordingly, the individual clinical, demographic and geographical pathway associations were considered exploratory

## Discussion

Parkinson’s disease presents as a recognizable clinical syndrome, yet its genetic architecture is distributed across interconnected cellular systems rather than confined to a single pathogenic route. The relative contribution of these systems may differ among populations because allele frequencies, founder effects and the distribution of monogenic and risk variants vary by ancestry. Nevertheless, South Asian populations remain substantially underrepresented in PD genetics, particularly in studies connecting genetic findings with biological pathways and clinical expression. This gap is especially relevant to juvenile- and young-onset disease, in which inherited determinants are more prominent. To our knowledge, the present study is among the earliest patient-level, pathway-resolved analyses integrating pathogenic/likely pathogenic variants, variants of uncertain significance and detailed clinical phenotypes in a South Asian PD cohort. Building upon a previously characterized^3,6,9^, multi-regional Indian cohort enriched for juvenile-, young- and early-onset PD, we moved beyond cataloguing individual genes to examine how genetic findings converge across cellular processes and whether this convergence relates to phenotypic variation.

Three findings define the study. First, the largest proportions of participants with reportable variants mapped to mitochondrial organization, autophagy–mitophagy and synaptic-vesicle trafficking, demonstrating extensive annotation-level convergence across cellular quality-control mechanisms. Second, the composition differed by evidential class: P/LP findings were concentrated in established early-onset genes, particularly *PRKN*, *PLA2G6* and *PINK1*, whereas lysosomal and cell-survival-related annotations were predominantly contributed by VUS involving *GBA1*, *VPS13C*, *LRRK2* and *FBXO7*. Third, among the 54 participants carrying a P/LP variant, those with additional VUS in distinct genes had an AAO comparable to P/LP-only carriers (30.5±10.4 versus 30.8±10.0 years; p=0.81). Family history was numerically more frequent in the P/LP-plus-VUS group but did not differ significantly (52.6% versus 31.4%; p=0.15). Multigene VUS-only carriers similarly showed no difference in AAO or family history compared with single-gene VUS carriers. Thus, co-occurring variants demonstrated biological convergence but were not associated with a detectable modifier phenotype in this cohort.

The pathway distribution is best understood as an interconnected mitochondrial– autophagic–lysosomal and vesicular quality-control network rather than as eight independent mechanisms. *PINK1* and *PRKN* connect mitochondrial damage sensing with mitophagic clearance, while *PLA2G6* influences mitochondrial membrane integrity, lipid homeostasis and autophagic processing. Their prominence is consistent with the early-onset composition of the cohort and with global studies identifying the PINK1– Parkin axis as a major contributor to recessive PD.^5,12–15^(Table–4) In a series of 1,587 patients selected for recessive or early-onset disease, biallelic *PRKN*, *PINK1* or *PARK7* variants were identified in 14.1%, with marked ethnic differences in the relative contribution of individual genes and *PRKN* remaining the most frequent cause overall.^16^. A subsequent multi-ancestry analysis demonstrated the broad global distribution of *PRKN* and observed that recessive *PRKN* variants were more frequent than causal *LRRK2* variants in Asian populations.^4^ The present findings extend these gene-level observations by showing how early-onset genes converge at the pathway level within an Indian clinical cohort.

**Table 4:** Selected international studies of genetic pathway involvement in Parkinson’s disease.

The lysosomal and trafficking-related findings provide a complementary layer of this architecture. Their representation was driven principally by VUS involving *GBA1*, *VPS13C*, *LRRK2*, *SPG11*, *ATP13A2* and related genes, placing several VUS-containing genes within a common lysosomal and endolysosomal framework, while not establishing functional impairment by the individual variants. This interpretation is consistent with case–control evidence extending lysosomal involvement beyond *GBA1*. Robak and colleagues demonstrated an aggregate excess of potentially damaging variants across 54 lysosomal-storage-disorder genes, with 56% of cases carrying at least one such variant and 21% carrying multiple alleles; the association remained significant after excluding *GBA1*.^5^ More recently, rare-variant analyses involving 8,267 patients and 68,208 controls implicated additional lysosomal processes, including sialylation, ganglioside metabolism and lysosomal proteolysis, with particularly relevant signals in early-onset PD.^17^ Synaptic vesicle trafficking further connects this network by regulating neurotransmitter release, endosomal recycling and delivery of proteins and organelles to degradative compartments. Thus, the mitochondrial, autophagic, lysosomal and synaptic categories identified here represent interacting components of neuronal homeostasis rather than competing explanations for disease.

The genetic composition of these pathways also illustrates how shared biological mechanisms may be populated by different variants across ancestries. No P/LP *LRRK2* variant was identified in this cohort, and all 26 *LRRK2* findings remained VUS. This differs from Ashkenazi Jewish, North African and Middle Eastern populations, in which p.G2019S contributes substantially to PD, but is compatible with the rarity of this causal variant in South and East Asian populations. Conversely, the prominence of *PRKN*, *PINK1* and *PLA2G6* reflects the early-onset and recessive architecture more frequently encountered in Asian PD series. The smaller iron-homeostasis/NBIA category, driven by *PLA2G6*, *ATP13A2*, *PANK2* and *WDR45*, was concentrated among younger patients and illustrates the biological continuum between juvenile PD and complex inherited parkinsonism. These findings suggest that populations may reach common pathogenic systems through different combinations of genes and variants—a distinction with implications for genetic diagnosis, biological modelling and therapeutic prioritization. Additional rare variants have been proposed as modifiers of genetically determined PD, although supporting evidence remains limited. Lubbe and colleagues reported that additional rare variants in Mendelian PD genes were more frequent among patients with an established primary genetic cause and were associated with younger onset. In contrast, the direct comparison in the present cohort did not identify an earlier AAO among P/LP-plus-additional-VUS carriers relative to P/LP-only carriers. Family history was numerically more frequent in the former group, but the difference was not statistically significant and the confidence interval was necessarily wide because of the small subgroup. Similarly, accumulation of VUS across multiple genes in the absence of a P/LP finding was not associated with AAO or familial clustering. Several combinations nevertheless involved genes mapping to interconnected mitochondrial, autophagic and lysosomal processes, including the PINK1–PRKN mitophagy axis. These combinations provide candidates for segregation, variant reclassification and functional investigation, but the present data do not establish oligogenic inheritance or clinical modification by additional VUS. The multiple lysosomal alleles reported by Robak and colleagues raised the possibility that rare variation across several components of cellular degradation pathways may contribute cumulatively to PD susceptibility. The present study extends the investigation of co-occurring variants to a clinically characterized Indian cohort. However, P/LP-plus-additional-VUS carriers did not have an earlier AAO than P/LP-only carriers, and the numerically higher frequency of family history did not reach statistical significance. Similarly, multigene VUS-only carriers did not differ from single-gene VUS carriers in AAO or familial clustering. Nevertheless, recurring combinations of *PRKN* or *PLA2G6* P/LP findings with VUS in genes involved in lysosomal and endolysosomal trafficking identify biologically plausible candidates for segregation, variant reclassification and functional studies. These observations do not establish oligogenic inheritance or a clinical modifier effect, but they define specific cross-pathway combinations in which such hypotheses can be tested.

The population setting adds further relevance. India encompasses substantial genetic diversity, long-standing endogamy and regional founder effects, making it unlikely that a single national variant spectrum captures its full PD architecture. The nominally greater representation of lysosomal and neuronal-survival annotations among patients of East Indian origin provides an initial indication that pathway profiles may vary even within India. Although this regional observation requires replication, it supports broader and more balanced recruitment across the country. At a global level, the findings also have implications for precision-medicine pipelines. Therapeutic programmes have concentrated particularly on *GBA1*- and *LRRK2*-associated biology, yet trial eligibility and target relevance depend on ancestry-specific variant spectra. The prominence of PINK1–Parkin, mitophagy and related early-onset pathways in this cohort suggests that therapeutic priorities derived predominantly from European populations may not fully represent South Asian disease architecture. VUS should not presently determine treatment or counselling, but pathway mapping can prioritize variants for functional resolution and help identify biological targets that merit investigation in diverse populations. The available categorical PRS analysis did not identify an association with pathway representation or clinical phenotype. This negative result neither establishes independence from polygenic background nor excludes rare–common variant interactions, particularly given the importance of ancestry calibration and the limitations of dichotomising PRS.

This study has several important strengths. It examines a large, multi-regional and deeply phenotyped Indian cohort derived from an established sequencing programme; integrates patient-level genetic, clinical and demographic information; separates P/LP from VUS-classified findings; and includes an internal comparison between P/LP-plus-VUS and VUS-only multigene states. These features allowed the analysis to progress from gene identification to clinically anchored pathway interpretation. The principal limitations define the next phase of investigation. Without ancestry-matched controls, the observed proportions represent pathway profiles within variant-positive patients rather than formal estimates of population-specific burden. Enrichment was conditioned by the predefined gene panels and available ontology annotations, and the pathways overlap extensively.. Most broader-panel findings were VUS, for which segregation, phase and functional evidence were generally unavailable The P/LP-plus-VUS subgroup was small, the same individuals contributed to several correlated pathway comparisons, and the exploratory symptom and regional associations did not survive correction for multiple testing nor exclude modest variant-specific effects. Family relatedness, incomplete segregation information and the absence of functional evidence further constrain causal interpretation of co-occurring variants. Replication in independent South Asian cohorts, formal pathway-burden testing against ancestry-matched controls and functional assessment of prioritized variants will be required to determine the generalizability and biological significance of the observed pathway convergence.

In conclusion, this study provides a pathway-resolved profile of reportable genetic variation in an Indian cohort enriched for juvenile-, young- and early-onset PD. The implicated genes converged principally across mitochondrial quality control, autophagy– mitophagy, lysosomal processing and vesicular trafficking, illustrating how multiple genetic findings can populate interconnected biological systems in an underrepresented population. Additional VUS in participants with established P/LP findings were not associated with earlier onset, emphasizing the distinction between gene-level biological convergence and demonstrated clinical modification. By integrating genetic classification, pathway annotation and detailed phenotype, this study establishes a framework for comparable analyses across global populations and for future validation using ancestry-matched controls, segregation and functional studies.

## Data Availability

De-identified data underlying this study may be made available by the GOPI YOPD Consortium upon reasonable request to the corresponding author, subject to approval by the consortiums data-access committee and applicable ethical, consent and data sharing requirements.

## Acknowledgements

The authors gratefully acknowledge all participants with Parkinson’s disease and their families for their time, trust and contribution to the Genetics of Parkinson’s Disease in India–Young-Onset Parkinson’s Disease (GOPI-YOPD) project. We also thank the investigators, clinical coordinators, laboratory and bioinformatics personnel, and staff at all participating centers whose collective efforts made this multicenter study possible. During preparation of this manuscript, the authors used OpenAI to assist with organization and language refinement.

---

**Supplementary Table 1.** Complete STRING functional-enrichment results for the pathogenic/likely pathogenic and variant-of-uncertain-significance gene sets

