## Supplementary material for "From genes to pathways: genetic convergence in early-onset Parkinson’s disease in India": Table-1

|  | **P/LP variant or VUS identified**  **(Total N=336)** | | | | | **No reportable P/LP variant or VUS identified**  **(Total N=332)** | | | | | **Grand Total (N=668)** |
| --- | --- | --- | --- | --- | --- | --- | --- | --- | --- | --- | --- |
| **Age group (yrs)** | **JOPD (<21)** | **YOPD**  **(21-40)** | **EOPD**  **(41-50)** | **ROPD**  **(>50)** | **Total** | **JOPD (<21)** | **YOPD (21-40)** | **EOPD**  **(41-50)** | **ROPD**  **(>50)** | **Total** | n/a |
| **Number of subjects** | 18 | 164 | 142 | 12 | 336 | 3 | 157 | 158 | 14 | 332 | 668 |
| **M:F** | 13:5 | 101:63 | 104:38 | 9:3 | 227:109 | 3:0 | 108:49 | 115:43 | 10:4 | 236:96 | 463:205 |
| **AAO, yrs, mean (range)** | 16.9  (11-20) | 33.6  (21-40) | 45.5 (  41-50) | 58.6  (51-66) | 38.6  (11-66) | 17.3  (14-20) | 34.1  (21-40) | 45.3  (41-50) | 57.0 (51-70) | 39.4  (11-70) | 39.4  (11-70) |
| **DOS, months, mean (range)** | 154.8  (17-360) | 104.4  (6-432) | 78.1  (2-276) | 47.0  (6-108) | 93.8  (2-432) | 220.0  (96-300) | 105.8  (2-360) | 88.7  (3-264) | 65.6  (8-200) | 97.2  (2-360) | 95.5  (2-432) |
| **Family History** | 6  (33.3%) | 39 (23.8%) | 26 (18.3%) | 10 (83.3%) | 81 (24.1%) | 1 (33.3%) | 25 (15.9%) | 21 (13.4%) | 11 (78.6%) | 58 (17.5%) | 139  (20.8%) |
| **Consanguinity** | 10 (55.6%) | 23 (14.3%) | 11  (7.8%) | 0  (0.0%) | 44 (13.3%) | 2 (66.7%) | 18 (12.0%) | 14 (8.9%) | 1  (7.1%) | 35 (10.8%) | 79  (12.0%) |
| **Geographical location** | | | | | | | | | | | |
| **North India** | 6  (33.3%) | 52 (31.7%) | 48 (33.8%) | 4  (33.3%) | 110 (32.7%) | 1  (33.3%) | 36  (22.9%) | 61 (38.6%) | 2  (14.3%) | 100 (30.1%) | 210  (31.4%) |
| **East India** | 3  (16.7%) | 36 (22.0%) | 33 (23.2%) | 3  (25.0%) | 75  (22.3%) | 0  (0.0%) | 32 (20.4%) | 28 (17.7%) | 0  (0.0%) | 60 (18.1%) | 135  (20.2%) |
| **South India** | 7  (38.9%) | 58 (35.4%) | 46 (32.4%) | 1  (8.3%) | 112  (33.3%) | 2 (66.7%) | 73 (46.5%) | 48 (30.4%) | 7 (50.0%) | 130  (39.2%) | 242  (36.2%) |
| **West India** | 1  (5.6%) | 14 (8.5%) | 13  (9.2%) | 4  (33.3%) | 32  (9.5%) | 0  (0.0%) | 11 (7.0%) | 19 (12.0%) | 4 (28.6%) | 34 (10.2%) | 66  (9.9%) |
| **Unknown region** | 1  (5.6%) | 4  (2.4%) | 2  (1.4%) | 0  (0.0%) | 7  (2.1%) | 0  (0.0%) | 5  (3.2%) | 2 (1.26%) | 1  (7.1%) | 8  (2.4%) | 15  (2.2%) |
| **Distribution of variant-class findings among participants with a reportable genetic finding** | | | | | | | | | | | |
| **P/LP-only** | 6  (33.3%) | 21  (12.8%) | 8  (5.6%) | 0  (0.0%) | 35  (10.4%) | n/a | n/a | n/a | n/a | n/a | n/a |
| **VUS-only** | 8  (44.4%) | 132 (80.5%) | 130 (91.5%) | 12 (100.0%) | 282 (83.9%) | n/a | n/a | n/a | n/a | n/a | n/a |
| **Dual-hit (1 P/LP + 1 VUS)** | 2 (11.1%) | 8  (4.9%) | 3  (2.1%) | 0  (0.0%) | 13 (3.9%) | n/a | n/a | n/a | n/a | n/a | n/a |
| **Triple-hit (1 P/LP + ≥2 VUS)** | 2  (11.1%) | 3  (1.8%) | 1  (0.7%) | 0  (0.0%) | 6  (1.8%) | n/a | n/a | n/a | n/a | n/a | n/a |

**Table -1 : Demographic, clinical and genetic characteristics of the study cohort, stratified by reportable genetic finding status and age-at-onset category**

*Values are n (%) unless otherwise specified. Percentages are calculated within the corresponding column using available-case denominators. The 26 participants with AAO >50 years were affected later-onset relatives recruited through qualifying families; 12 had a reportable VUS and 14 had no reportable P/LP variant or VUS. AAO, age at onset; DOS, duration of symptoms; JOPD, juvenile-onset Parkinson’s disease; YOPD, young-onset Parkinson’s disease; EOPD, early-onset Parkinson’s disease; ROPD, Regular-onset Parkinson’s disease; P/LP, pathogenic or likely pathogenic; VUS, variant of uncertain significance. n/a: Not applicable*
