## Supplementary material for "From genes to pathways: genetic convergence in early-onset Parkinson’s disease in India": Table-2

**Table 2. Patient-level representation and clinical characteristics across eight collated gene-based pathway categories**

| **Characteristic** | **Mitochondrial organization** | **Autophagy/ mitophagy** | **Lysosomal function** | **Dopamine regulation** | **Synaptic-vesicle trafficking** | **Regulation of neuron apoptotic process** | **Oxidative-stress response** | **Iron homeostasis/ NBIA-related processes** |
| --- | --- | --- | --- | --- | --- | --- | --- | --- |
| **Sample size and gender** | | | | | | | | |
| Participants, n (% of cohort; N=336) | 247 (73.5) | 228 (67.9) | 143 (42.6) | 139 (41.4) | 201 (59.8) | 201 (59.8) | 158 (47.0) | 42 (12.5) |
| Male sex, n (%) | 167 (67.6) | 152 (66.7) | 95 (66.4) | 95 (68.3) | 139 (69.2) | 137 (68.2) | 108 (68.4) | 32 (76.2) |
| High PRS, n/N (%) | 59/231 (25.5) | 55/212 (25.9) | 36/132 (27.3) | 29/129 (22.5) | 52/191 (27.2) | 55/185 (29.7) | 39/147 (26.5) | 10/39 (25.6) |
| **Age and disease course** | | | | | | | | |
| Age at onset, mean ± SD, years | 37.8 ± 9.8 | 37.2 ± 9.7 | 39.1 ± 8.4 | 37.3 ± 10.6 | 38.3 ± 10.2 | 39.2 ± 9.1 | 36.8 ± 10.5 | 34.8 ± 12.4 |
| Duration of symptoms, mean ± SD, months | 95 ± 78 | 98 ± 80 | 86 ± 70 | 94 ± 87 | 98 ± 81 | 90 ± 71 | 95 ± 86 | 88 ± 83 |
| JOPD (≤20 years), n (%) | 17 (6.9) | 18 (7.9) | 7 (4.9) | 13 (9.4) | 13 (6.5) | 11 (5.5) | 16 (10.1) | 7 (16.7) |
| YOPD (21–40 years), n (%) | 121 (49.0) | 113 (49.6) | 67 (46.9) | 64 (46.0) | 99 (49.3) | 89 (44.3) | 72 (45.6) | 20 (47.6) |
| EOPD (41–50 years), n (%) | 102 (41.3) | 93 (40.8) | 67 (46.9) | 56 (40.3) | 77 (38.3) | 95 (47.3) | 67 (42.4) | 13 (31.0) |
| ROPD (>50 years), n (%) | 7 (2.8) | 4 (1.8) | 2 (1.4) | 6 (4.3) | 12 (6.0) | 6 (3.0) | 3 (1.9) | 2 (4.8) |
| **Family history and consanguinity** | | | | | | | | |
| Family history of PD, n/N (%) | 64/247 (25.9) | 60/228 (26.3) | 37/143 (25.9) | 39/139 (28.1) | 51/201 (25.4) | 46/201 (22.9) | 41/158 (25.9) | 14/42 (33.3) |
| Consanguinity, n/N (%) | 33/243 (13.6) | 29/224 (12.9) | 13/139 (9.4) | 20/138 (14.5) | 31/198 (15.7) | 16/197 (8.1) | 25/157 (15.9) | 9/40 (22.5) |
| **Region of family origin** | | | | | | | | |
| North India, n (%) | 81 (32.8) | 72 (31.6) | 43 (30.1) | 39 (28.1) | 64 (31.8) | 60 (29.9) | 43 (27.2) | 16 (38.1) |
| South India, n (%) | 82 (33.2) | 73 (32.0) | 41 (28.7) | 50 (36.0) | 68 (33.8) | 60 (29.9) | 56 (35.4) | 16 (38.1) |
| East India, n (%) | 53 (21.5) | 54 (23.7) | 44 (30.8) | 28 (20.1) | 46 (22.9) | 53 (26.4) | 34 (21.5) | 9 (21.4) |
| West India, n (%) | 25 (10.1) | 23 (10.1) | 12 (8.4) | 16 (11.5) | 17 (8.5) | 24 (11.9) | 20 (12.7) | 1 (2.4) |
| Other/unknown region, n (%) | 6 (2.4) | 6 (2.6) | 3 (2.1) | 6 (4.3) | 6 (3.0) | 4 (2.0) | 5 (3.2) | 0 (0.0) |

***Table 2 (continued)***

| **Characteristic** | **Mitochondrial organization** | **Autophagy/ mitophagy** | **Lysosomal function** | **Dopamine regulation** | **Synaptic-vesicle trafficking** | **Regulation of neuron apoptotic process** | **Oxidative-stress response** | **Iron homeostasis/ NBIA-related processes** |
| --- | --- | --- | --- | --- | --- | --- | --- | --- |
| **Dyskinesia** | | | | | | | | |
| Developed dyskinesia, n/N (%) | 82/244 (33.6) | 76/225 (33.8) | 44/140 (31.4) | 39/138 (28.3) | 64/197 (32.5) | 62/199 (31.2) | 46/157 (29.3) | 12/42 (28.6) |
| Time to dyskinesia, median (IQR), years | 5.0 (2.0–10.2) | 5.5 (2.8–12.0) | 6.0 (4.0–12.0) | 7.0 (3.5–10.5) | 5.0 (2.5–10.5) | 5.0 (2.0–7.0) | 5.0 (2.0–9.5) | 2.0 (1.8–10.0) |
| **Motor features at presentation** | | | | | | | | |
| Rest tremor, n/N (%) | 186/244 (76.2) | 170/225 (75.6) | 104/143 (72.7) | 99/136 (72.8) | 145/199 (72.9) | 145/197 (73.6) | 119/155 (76.8) | 33/41 (80.5) |
| Rigidity, n/N (%) | 191/243 (78.6) | 176/224 (78.6) | 108/139 (77.7) | 110/139 (79.1) | 162/200 (81.0) | 154/198 (77.8) | 128/158 (81.0) | 29/41 (70.7) |
| Bradykinesia, n/N (%) | 220/246 (89.4) | 209/227 (92.1) | 130/141 (92.2) | 123/137 (89.8) | 179/199 (89.9) | 184/200 (92.0) | 143/157 (91.1) | 36/42 (85.7) |
| Gait impairment, n/N (%) | 140/243 (57.6) | 129/224 (57.6) | 76/141 (53.9) | 78/137 (56.9) | 115/197 (58.4) | 111/197 (56.3) | 90/156 (57.7) | 28/41 (68.3) |
| Dystonia, n/N (%) | 50/236 (21.2) | 48/220 (21.8) | 27/137 (19.7) | 33/133 (24.8) | 51/194 (26.3) | 40/190 (21.1) | 38/154 (24.7) | 12/41 (29.3) |
| Asymmetric motor onset, n/N (%) | 228/241 (94.6) | 210/222 (94.6) | 135/139 (97.1) | 126/135 (93.3) | 188/197 (95.4) | 190/197 (96.4) | 146/155 (94.2) | 37/41 (90.2) |
| **Non-motor features during the course of illness** | | | | | | | | |
| Apathy, n/N (%) | 75/245 (30.6) | 73/226 (32.3) | 50/143 (35.0) | 44/139 (31.7) | 66/201 (32.8) | 70/198 (35.4) | 53/157 (33.8) | 12/42 (28.6) |
| Anxiety, n/N (%) | 109/246 (44.3) | 105/228 (46.1) | 67/143 (46.9) | 59/138 (42.8) | 89/200 (44.5) | 98/201 (48.8) | 73/158 (46.2) | 23/42 (54.8) |
| Panic attacks, n/N (%) | 39/243 (16.0) | 37/225 (16.4) | 23/141 (16.3) | 23/136 (16.9) | 35/197 (17.8) | 30/198 (15.2) | 22/155 (14.2) | 11/40 (27.5) |
| Depression, n/N (%) | 112/246 (45.5) | 103/228 (45.2) | 66/143 (46.2) | 68/139 (48.9) | 101/200 (50.5) | 94/201 (46.8) | 73/158 (46.2) | 20/41 (48.8) |
| Hallucinations or delusions, n/N (%) | 25/245 (10.2) | 26/227 (11.5) | 16/143 (11.2) | 13/138 (9.4) | 22/199 (11.1) | 19/200 (9.5) | 15/157 (9.6) | 5/41 (12.2) |
| Memory/cognitive symptoms, n/N (%) | 49/245 (20.0) | 44/227 (19.4) | 27/143 (18.9) | 30/138 (21.7) | 49/199 (24.6) | 44/200 (22.0) | 34/157 (21.7) | 11/41 (26.8) |
| Light-headedness/dizziness, n/N (%) | 18/245 (7.3) | 15/227 (6.6) | 10/143 (7.0) | 9/139 (6.5) | 20/200 (10.0) | 13/200 (6.5) | 11/158 (7.0) | 3/41 (7.3) |
| Orthostatic hypotension, n/N (%) | 9/245 (3.7) | 9/227 (4.0) | 6/143 (4.2) | 6/139 (4.3) | 11/200 (5.5) | 9/200 (4.5) | 7/157 (4.5) | 4/41 (9.8) |
| Constipation, n/N (%) | 100/246 (40.7) | 94/228 (41.2) | 65/143 (45.5) | 45/139 (32.4) | 71/198 (35.9) | 89/201 (44.3) | 52/158 (32.9) | 17/41 (41.5) |

Values are n (%) unless otherwise stated. Pathway categories were collated from significant functional-enrichment terms identified separately in the P/LP and VUS gene sets. Categories are non-mutually exclusive: participants may be represented in more than one category, and pathway percentages therefore do not sum to 100%. Clinical percentages use available-case denominators. The exact STRING terms, ontology identifiers, contributing genes, observed and expected gene counts, enrichment strengths, FDR values and category-consolidation rules are provided in the Supplementary Methods and Tables. Nominal and multiple-testing-adjusted pathway–phenotype comparisons are reported in the Supplementary Tables. EOPD, early-onset Parkinson’s disease; FDR, false-discovery rate; IQR, interquartile range; JOPD, juvenile-onset Parkinson’s disease; ROPD, Regular-onset Parkinson’s disease; NBIA, neurodegeneration with brain iron accumulation; PD, Parkinson’s disease; P/LP, pathogenic or likely pathogenic; PRS, polygenic risk score; SD, standard deviation; VUS, variant of uncertain significance; YOPD, young-onset Parkinson’s disease.
