## Supplementary material for "From genes to pathways: genetic convergence in early-onset Parkinson’s disease in India": Table-3

Table - 3: Comparison of clinical, demographic and genetic characteristics between participants carrying a pathogenic/likely pathogenic variant alone and those carrying a pathogenic/likely pathogenic variant plus one or more variants of uncertain significance in additional genes.

| Variable | PLP-only | PLP + 1 VUS gene (dual-hit) | PLP + 2 VUS genes (triple-hit) | PLP + Any additional VUS gene |
| --- | --- | --- | --- | --- |
| N | 35 | 13 | 6 | 19 |
| M:F | 23:12 | 8:5 | 4:2 | 12:7 |
| AAO (mean yr ± SD) | 30.8 ± 10.0 | 31.3 ± 10.8 | 28.7 ± 10.4 | 30.5 ± 10.4 |
| DOS (months ± SD) | 136.8 ± 109.2 | 90.5 ± 75.7 | 115.0 ± 128.3 | 98.7 ± 93.2 |
| JOPD (n) | 6 | 2 | 2 | 4 |
| YOPD (n) | 21 | 8 | 3 | 11 |
| EOPD (n) | 8 | 3 | 1 | 4 |
| LOPD (n) | 0 | 0 | 0 | 0 |
| FH, n/N (%) | 11/35 (31.4%) | 7/13 (53.8%) | 3/6 (50.0%) | 10/19 (52.6%) |
| Consanguinity, n/N (%) | 13/35 (37.1%) | 3/13 (23.1%) | 2/6 (33.3%) | 5/19 (26.3%) |
| North India (n) | 8 | 8 | 1 | 9 |
| East India (n) | 4 | 0 | 0 | 0 |
| South India (n) | 20 | 3 | 4 | 7 |
| West India (n) | 1 | 2 | 0 | 2 |
| Other/Bangladesh (n) | 2 | 0 | 1 | 1 |
| High PRS, n/N (if available) | 5/33 (15.2%) | 5/12 (41.7%) | 0/5 (0.0%) | 5/17 (29.4%) |
| Primary PLP genes | PRKN n=21, PINK1 n=4, CHCHD2 n=2, PLA2G6 n=2, GCH1 n=1, MAPT n=1, PRRT2 n=1, SYNJ1 n=1, VPS13C n=1, WDR45 n=1 | PRKN n=4, PLA2G6 n=3, CHCHD2 n=2, ATP13A2 n=1, MAPT n=1, PINK1 n=1, VPS13C n=1 | PRKN n=4, PLA2G6 n=2 | PRKN n=8, PLA2G6 n=5, CHCHD2 n=2, ATP13A2 n=1, MAPT n=1, PINK1 n=1, VPS13C n=1 |

No significant correlations were noted between PLP only vs PLP with additional variant
