## Supplementary material for "From genes to pathways: genetic convergence in early-onset Parkinson’s disease in India": Table-4

**Table – 4 : Selected international studies of genetic pathway involvement in Parkinson’s disease**

| **Study** | **Population and sample** | **Genetic approach** | **Principal pathway finding** | **Relationship to the present study** |
| --- | --- | --- | --- | --- |
| Lesnick et al., 2007 | USA; 443 PD–sibling-control pairs | GWAS SNP-set analysis | Axon-guidance signalling | Early demonstration of pathway-level convergence among distributed common variants |
| Holmans et al., 2013 | Europe/USA; 12,386 cases and 21,026 controls | GWAS pathway-enrichment analysis | Antigen processing and immune pathways | Large-scale common-variant pathway analysis in predominantly European populations |
| Robak et al., 2017 | Predominantly Europe/USA; discovery cohort of 1,156 cases and 1,679 controls, with two replication cohorts | WES rare-variant burden testing across 54 lysosomal genes | Lysosomal-storage-disorder gene burden | Methodological benchmark for formal case–control rare-variant pathway burden |
| Bandres-Ciga et al., 2020 | Predominantly European ancestry; 26,035 cases and 403,190 controls | MAGMA analysis of a curated trafficking gene set | Endocytic membrane trafficking | Supports distributed common-variant enrichment within vesicular and endosomal trafficking |
| Zhao et al., 2021 | China; 3,879 patients and 2,931 controls | WES/WGS and SKAT-O across 69 lysosomal genes | Lysosomal rare-variant burden, particularly in familial and early-onset PD | Provides an ancestry-specific Asian comparator, although restricted to one predefined pathway |
| Present study | India; parent cohort n=668, including 336 participants with reportable P/LP or VUS findings | Separate enrichment of 11-gene P/LP and 40-gene VUS sets, followed by patient-level pathway mapping and clinical correlation | Convergence across eight overlapping biological domains | Extends clinically integrated, multipathway analysis to an underrepresented South Asian early-onset cohort; evaluates pathway representation rather than case–control burden |

Studies were selected to illustrate major pathway-based genetic approaches in PD and do not constitute a systematic review. Direct comparison of pathway frequencies is limited by differences in ancestry, study design, variant classes, gene-set definitions, reference populations and statistical methods.
